# Clinician Readiness to Adopt A.I. for Critical Care Prioritisation

**DOI:** 10.1101/2021.02.11.21251604

**Authors:** Georgina Kennedy, Blanca Gallego

## Abstract

There is a wide chasm between what has been shown to be feasible in the application of artificial intelligence to data from the electronic medical record, and what is currently available. The reasons for this are complex and understudied, and vary across technical, ethical and sociocultural domains. This work addresses the gap in the literature for studies that determine the readiness of clinical end-users to adopt such tools and the way in which they are perceived to affect clinical practice itself.

In this study, we present a novel, credible AI system for predicting in-patient deterioration to likely end users. We gauge their readiness to adopt this technology using a modified version of the technology adoption model.

Users are found to be moderately positive towards the potential introduction of this technology in their workflow, although they demonstrate particular concern for the appropriateness of the clinical setting into which it is deployed.

## 1. Introduction

### 1.1. Background

Within the recent proliferation of reviews and summaries of the state of artificial intelligence (AI) in clinical settings [1,2,3,4,5,6], there is evidence of successful translation of AI research into actual clinical practice for the analysis of images [7,8,9,10,11]^2^. However, when it comes to the domain of AI decision support based on data from the electronic medical record (EMR), despite much interest and a number of viable models [12,13,14], there are very few signs of mature real-world implementations of such systems.

Asides from the technical and procedural challenges of data harmonisation and integration, model generalisability and ethical safeguards [15,3,16], and the high bar of achieving approval of software as a medical device [17], there is of course also a human and cultural barrier to entry that must be crossed in order to successfully implement these tools in practice. Despite this, there is no prior work to our knowledge that presents a credible EMR-based AI decision support system to likely clinician users to assess their opinion of its suitability within their daily work.

The work presented in [18] describes an AI system for predicting risk of in-patient deterioration, targeted to the specific use-case of prioritising the work of the critical care outreach team. This system uses only real-time available data and is designed with the goal of being integratable within the EMR.

### 1.2. Objective

The aim of this study is to assess the readiness of intensive care clinicians and leaders to adopt an AI-based decision support system for the prioritisation of patients at risk of deterioration on the wards.

The critical care outreach workflow is a reasonable one in which to pilot this emergent technology because it is a challenging and fast-paced role that requires rapidly updating awareness of events across the whole hospital. It is therefore a natural fit for any tool that can reliably synthesise a large amount of real-time data to augment clinical judgement. In addition, it already typically relies on track-and-trigger early warning systems (EWS) [19], and therefore the progression to what is effectively a risk model based on a broader selection of data is not as great a leap as it may be in other contexts.

We propose that this is a necessary next step towards completion of an appropriate impact study, as the complexity of such an implementation (even in pilot phase) cannot be understated. A full and theoretically grounded account of stakeholder readiness and understanding of potential pain points will be a powerful tool in navigating an intricately balanced set of clinical, cultural and technological priorities required for its successful execution.

### 1.3. Prior Work

The original technology acceptance model (TAM) [20] and its derivatives [21,22] have been used to understand barriers to technology adoption in the context of many disruptive technologies such as wireless internet [23], e-commerce [24], and personal computing [25], and importantly have been demonstrated for healthcare applications such as telemedicine [26], electronic medical records [27] and mobile healthcare systems [28].

The TAM framework explains the behavioural intention of an individual to use a new technology as a factor of their attitude towards its use. Their attitude towards a technology is affected directly by the measures of its *perceived usefulness* (PU) and *perceived ease of use* (PEOU). The premise of this model is that an individual’s attitude to using a system is a good predictor of their behavioural intention to use it (BI), and in turn, their actual eventual use.

The review papers [29,30] found that whilst the TAM has general capacity to predict technology acceptance in a clinical or healthcare setting, it is necessary to include additional context-specific explanatory variables.

We reviewed the literature for expanded versions of the TAM that had been validated in a healthcare setting [31,32] in order to adopt a minimal set of relevant additional variables for this use-case. The proposed additional variables were kept to a minimal set of demographic questions in order to reduce the burden of response on the target subjects, in particular given the timing of the study during the outbreak of COVID-19 and the extraordinary pressure on intensive care teams during this time. It was deemed unlikely that within the highly targeted (and therefore small) population of probable users, sufficient responses could be gathered to validate any new constructs, so added variables were restricted to modulating factors only.

### 1.4. Research Model and Hypotheses

The TAM is comprised of the hypotheses summarised as H1–5 in Table 1. We further expanded the model with three additional context-specific hypotheses. We do not include the TAM2 second and third order factor antecedents of perceived usefulness [21] due to the challenge of meaningfully capturing these factors for a proposed (not implemented) system.

**Table 1.** Hypotheses proposed by the TAM (H1-5), extended with H3b and H6-8 which are specific to this context

|  | Hypothesis | As Equation |
| --- | --- | --- |
| H1 | Behavioural intent predicts actual use | BI→AU |
| H2 | Attitude to use explains behavioural intent | ATT→BI |
| H3 | Perceived usefulness explains attitude | PU→ATT |
| H4 | Perceived ease of use explains attitude | PEOU→ATT |
| H5 | Perceived ease of use explains perceived usefulness | PEOU→PU |
| H3b | Perceived usefulness will be the most important model factor in driving attitude | PU→ATT > PEOU→ATT |
| H6 | Clinical role modulates perceived usefulness | ROLE~PU |
| H7 | Time devoted to task modulates perceived usefulness | TIME~PU |
| H8 | Experience modulates perceived usefulness | EXP~PU |

**Table 2.**
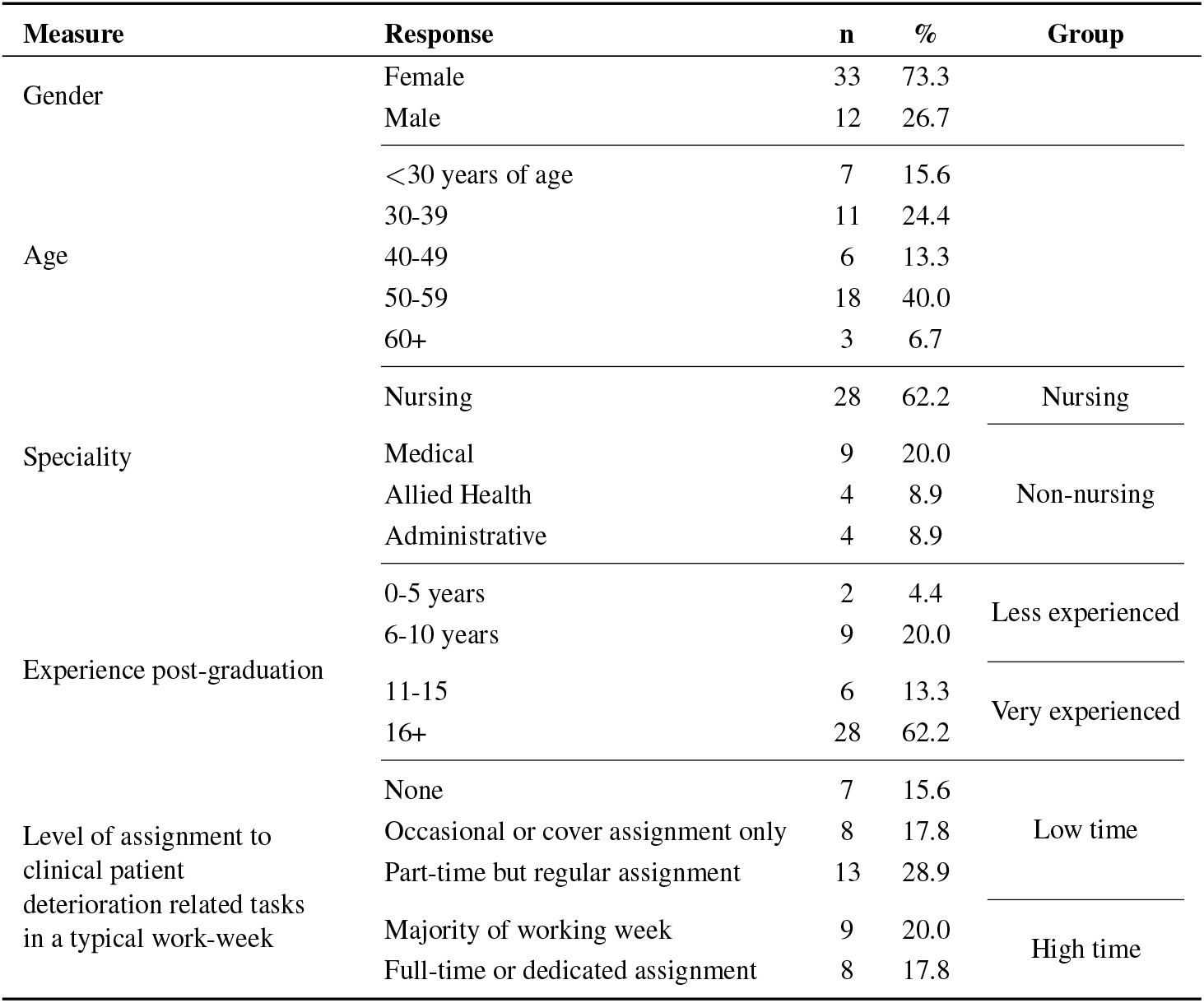
Respondent Demography

It is hypothesised that the perception of usefulness will be the most significant factor in the successful adoption of this technology (*H3b*), as the capacity for a model to improve the workflow itself is a critical factor for clinician buy-in. This is particularly true in this early stage, where it is necessary to confirm whether the model is even addressing a real need [33]. This hypothesis is supported by the observation made by [30] that in 100% of reviewed healthcare TAM studies, the PU *→* BI relationship was significant, compared to just over half of reviewed studies finding a statistically significant PEOU*→* BI relationship.

This study expands the existing TAM hypotheses with three additional modulating factors:

- The perception of usefulness will be modulated by how well a potential user can envisage such a system supporting their own workflow, and therefore will be dependent upon their role (*H6*)
- A user who spends most of their working week assigned to relevant patient deterioration tasks will see greater value in automated data synthesis to support the workflow, increasing their perception of its usefulness (*H7*)
- Senior patient deterioration staff may be more separated from the day to day challenges imposed by interaction with clinical information management systems, and therefore it will affect their view of how practical an AI system would be in practice (*H8*)

## 2. Methods

### 2.1. Data Collection

The questionnaire consisted of two parts. The first section captured variables that described the subject’s role, experience and demography. After responding to the demography questions, subjects were asked to review a prototype user interface, alongside descriptions of the tasks being performed (reproduced in the Appendix). In addition, some high-level summary information was provided on the development of the predictive model itself (data sources, mode of operation and accuracy — as described in [18]).

The main portion of the questionnaire was defined by adapting the measures in the original validated model [21] to the target context. A single free-text response was also captured that allowed respondents to make any additional comments or suggestions that they felt were pertinent to the potential roll-out of such a tool. Finally, in order to identify disingenuous responses, a question to test malingering with a trivially correct response was included.

The survey was distributed by email and completed online using the Qualtrics survey platform. All TAM measures were captured as a 7 item Likert scale from 1 = Strongly Disagree to 7 = Strongly Agree.

### 2.2. Population

This questionnaire was distributed to members of the New South Wales Deteriorating Patient Advisory Group (NSW-DPAG) (n *∼* 170), which is made up of highly engaged critical care staff, including nurses, physicians and administrators across the public healthcare system. This represents the key decision-making group whose advocacy and leadership would be a necessary prerequisite for a successful pilot implementation of this system within their respective institutions.

### 2.3. Data Analysis

A structural equation modeling approach was used for this analysis, based on the lavaan library [34] in R.

## 3. Results

### 3.1. Descriptive Statistics

The questionnaire was sent out to the NSW-DPAG member email list in July of 2020 and was open for a period of one month. 59 responses were captured, giving a 34.7% response rate. 14 response sets were removed for not proceeding beyond review of the proposed application. None of the remaining sets provided an incorrect or missing response on the malingering item, and therefore 45 response sets were retained for analysis. 91% of the remaining respondents (41) completed all measures, giving an overall completion rate of 96% for the analysis data.

Overall the respondents were highly experienced, with the majority coming from a nursing background (60%). This is expected within the included group, and accurately represents the key decision makers who would be responsible for overseeing the implementation of such a system. Managerial levels are over-represented in this sample, however 67% had at least some regular assignment to relevant clinical tasks and therefore fit the profile of an expected end-user as well.

### 3.2. Model Validation

#### 3.2.1. Construct Reliability

Re-validation of the model constructs was necessary, due to their modification to fit the research context.

After applying a mean imputation strategy, we cannot reject the null hypothesis of data non-normality (tested with [35] due to small sample size), and therefore are unable to use maximum likelihood estimation to fit our model. A robust unweighted least squares strategy [36] was thus used instead.

Despite a moderate response rate, the final number of responses was insufficient to assess the reliability of the original 4 factors comprising the TAM (model non-convergence). We therefore follow the lead of the updated TAM2 model [21] in merging the BI and ATT latent factors. In doing this, some of the nuance between a potential user believing the system overall is a good idea and it translating into their actual intention to use it is lost, but a simpler and more robust model is produced, which can withstand analysis under a small data set. The high composite reliability for this merged factor (higher than the average CR for each individual construct in [37]) also supports this action.

**Figure 1.**
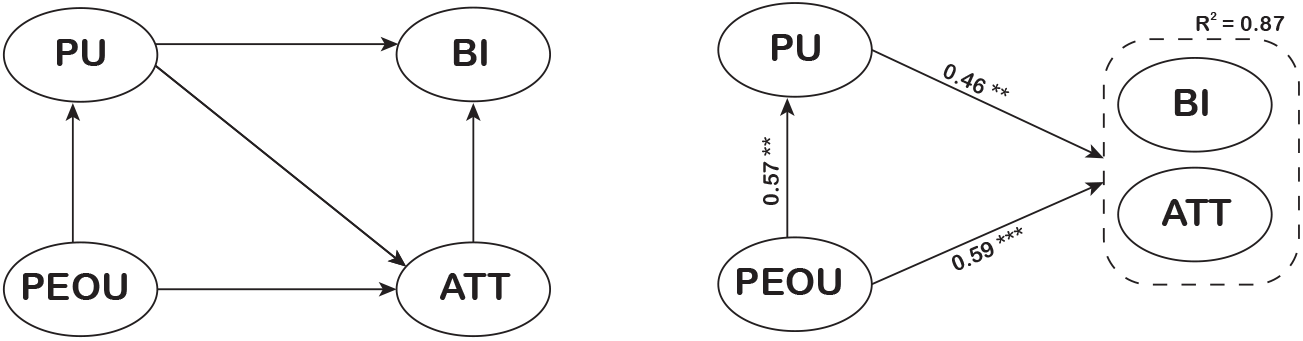
Original TAM (left) with adjustments for this context (right) (standardised factor weightings)

The model was further simplified by removing one PU item (PU3) and one PEOU item (PEOU4) due to their high degree of collinearity with other items in their respective constructs.

In all instances, the resultant construct composite reliability (CR) meets the 0.8 standard for generally acceptable reliability [38,39] (see Table 3). The average variance extracted (AVE) for each construct also exceeds the threshold of 0.5, required as per [40] to ensure that variance due to measurement error does not exceed the variance of the construct itself.

**Table 3.** Confirmatory Factor Analysis

| Construct | Items | Factor Loading | AVE | Composite Reliability |
| --- | --- | --- | --- | --- |
| PU | PU1 | 0.970 | 0.885 | 0.938 |
|  | PU2 | 0.910 |  |  |
| PEOU | PEOU1 | 0.915 | 0.607 | 0.820 |
|  | PEOU2 | 0.678 |  |  |
|  | PEOU3 | 0.723 |  |  |
| BI/ATT | BI1 | 0.847 | 0.694 | 0.901 |
|  | BI2 | 0.862 |  |  |
|  | ATT1 | 0.768 |  |  |
|  | ATT2 | 0.853 |  |  |

#### 3.2.2. Model Fit and Assessment of Hypotheses

The Satorra-Bentler scaled chi-squared test statistic allows the generation of goodness of fit indices that do not make any assumptions about the normality of the underlying data [41]. Applying this method, we do not reject the null hypothesis of good model fit (p=0.281). Other model fit indices were also indicative of good fit: CFI = 0.989, TLI = 0.983, SRMR = 0.058.

Most importantly, those relationships that have been retained in the simplified model are theoretically grounded. We therefore accept PU and PEOU as antecedents of a user’s intention to use this system, and accept that the measures in this survey tool are sufficient to quantify PU, PEOU and BI constructs for these exploratory purposes.

Given the hypothetical nature of this target system, it is naturally impossible to test *H1* in advance of any meaningful pilot, although it is retained here for completeness, and as per section 3.2.1, we had insufficient statistical power to test *H2*.

Consistent with the existing literature, both perceived usefulness and perceived ease of use were significant determinants of behavioural intent to use (Table 4), and therefore *H3* and *H4* were supported. Perceived ease of use was also a significant antecedent of perceived usefulness, supporting *H5*.

**Table 4.**
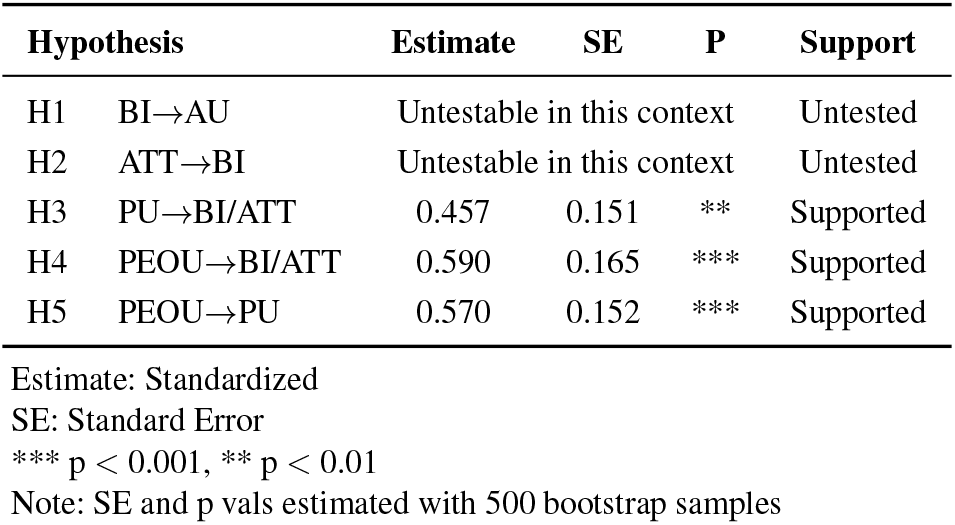
Support for Hypotheses (TAM)

| Hypothesis | Estimate | SE | P | Support |
| --- | --- | --- | --- | --- |
| H1 BI→AU | Untestable in this context |  |  | Untested |
| H2 ATT→BI | Untestable in this context |  |  | Untested |
| H3 PU→BI/ATT | 0.457 | 0.151 | ** | Supported |
| H4 PEOU→BI/ATT | 0.590 | 0.165 | *** | Supported |
| H5 PEOU→PU | 0.570 | 0.152 | *** | Supported |

Estimate: Standardized
SE: Standard Error
\*\*\* $p < 0.001$ , \*\* $p < 0.01$
Note: SE and $p$ vals estimated with 500 bootstrap samples

The standardized estimates of the relationship between PU and BI/ATT and PEOU and BI/ATT are similar, and in fact PEOU is found to be somewhat higher in weighting. This is the opposite effect as proposed by *H3b*, so it is not supported.

#### 3.2.3. Weighted Results

Table 5 reports the mean and standard deviation for each factor (weighted and un-weighted). The ratio of weighted mean to the theoretical weighted maximum is above 0.5 for each construct, showing that overall the potential users rated the proposed application as relatively useful and relatively easy to use, leading to an overall positive attitude and intention to use.

**Table 5.** Weighted factors

| Construct<br>(items) | Unweighted |  | Weighted |  | Range - actual<br>(theoretical) | WM/TM |
| --- | --- | --- | --- | --- | --- | --- |
|  | Mean | SD | Mean | SD |  |  |
| PU (2) | 10.42 | 2.32 | 9.79 | 2.18 | 2.85-13.16 (1.88-13.16) | 0.74 |
| PEOU (3) | 15.23 | 3.30 | 11.68 | 2.59 | 5.94-16.21 (2.32-16.21) | 0.72 |
| BI/ATT (4) | 20.62 | 4.32 | 17.14 | 3.61 | 6.66-22.45 (3.33-23.31) | 0.76 |

Weightings: Standardized
WM/TM - ratio of weighted mean to theoretical maximum for this construct

Both PU and PEOU saw ceiling effects, where at least one respondent answered maximally positively (strongly agree) across the whole measure. No respondent answered maximally negatively (strongly disagree) across any measure. This is particularly true for PEOU, which had a minimum unweighted score of 8 out of a possible 21. The ranges of responses overall, however, were quite wide.

### 3.3. Multi-group Analysis

#### 3.3.1. Model Invariance

To assess hypotheses that predict mediation of relationships between groups (H6, H7, H8), measurement invariance must first be confirmed. We first establish a baseline model to confirm the factor-loading pattern between groups (configural model), where the factor structure is the same, but all other elements of the model are allowed to vary freely between groups. This is compared to models where invariance is enforced for (1) factor loadings (metric model), (2) both factor loadings and model intercepts (scalar model) and (3) factor loadings, model intercepts and item variances (strict model) [42].

When comparing the *Nursing vs. Non-Nursing* and *High time vs. Low time* intergroup relationships, note that in each case, a single negative variance was produced, which is potentially indicative of model misspecification. In this case, however, it is likely to be due to the low group sizes, as in each instance the estimate plus the respective standard error was positive [43]. Conversely, when comparing the *Highly Experienced* group to the *Moderately or Less Experienced group*, more samples are required in order to define a fully-specified model, so it was not possible to test H8.

Table 6 shows that there is good model fit for the configural, metric and scalar models, and therefore latent mean analysis is valid, as the latent factors can be assumed to be invariant in configuration, factor loadings and scale.

**Table 6.**
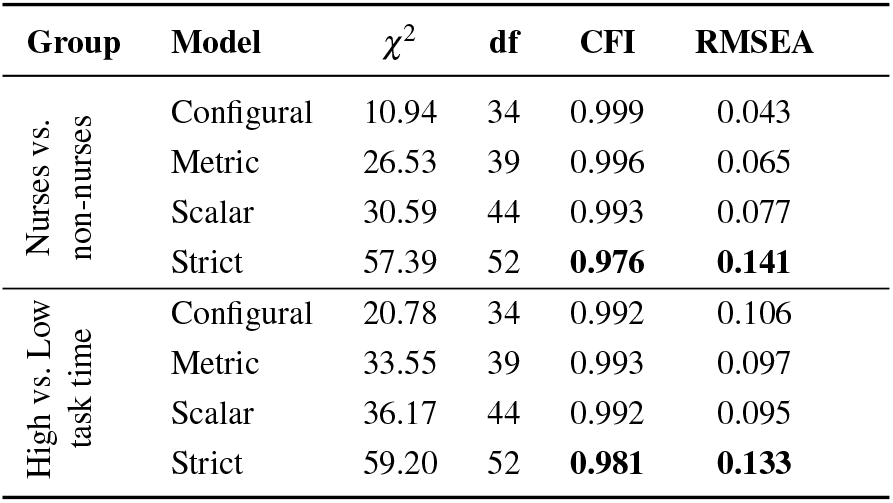
Inter-group model comparisons

For the strict model, there is a change in the CFI that exceeds the threshold of −0.010 combined with an increase in RMSEA by more than 0.010, as suggested in [44]. The strict model change in fit implies that the variance in responses may differ across groups, despite the overall model structure remaining valid. To estimate the differences in latent-factor means across groups, we constrain the factor mean in a reference group to zero, and then estimate the mean in the comparison group to produce the difference in factor mean [45]. This was done individually for each latent factor in the model and is reported in Table 7.

**Table 7.**
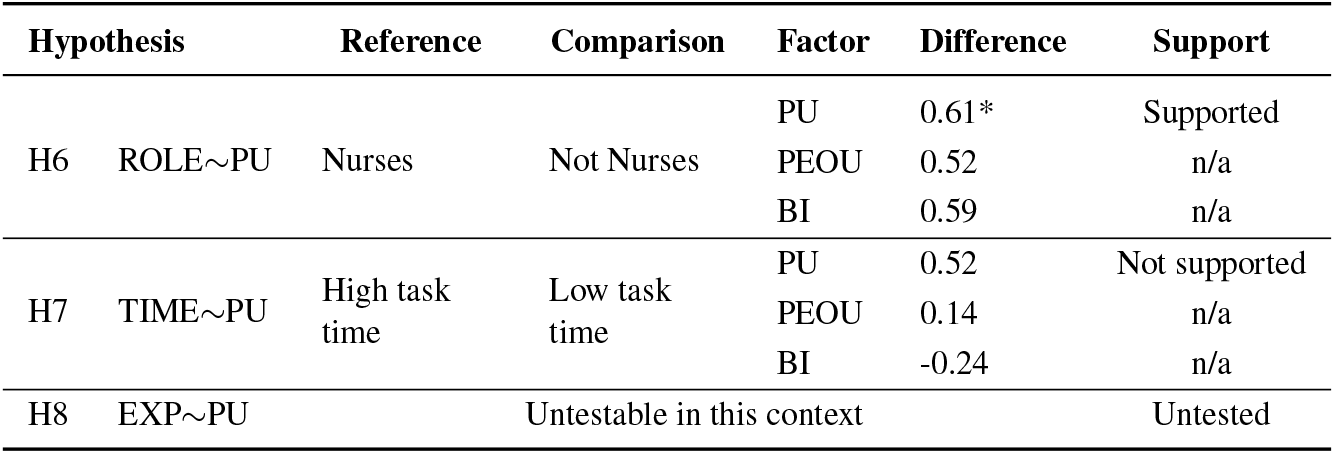
Hypothesis testing for modulating factors

**Table 8.** Demography Measures

| Question | Responses |
| --- | --- |
| Age | Under 30 years of age<br>30-39 years<br>40-49 years<br>50-59 years<br>60+ years<br>Prefer not to respond |
| Gender | Male<br>Female<br>Prefer not to respond<br>Other |
| Level of assignment to clinical patient deterioration related tasks (including outreach and emergency response) in a typical work-week | None<br>Occasional or cover assignment only<br>Part-time but regular assignment<br>Majority of working week<br>Full-time or dedicated assignment |
| Role | Nursing<br>Medical<br>Administrative<br>Other |
| Years of experience post-graduation | 0-5<br>6-10<br>11-15<br>16+ |

**Table 9.** TAM Measures

| Measure | Questions |
| --- | --- |
| ATT | ATT1 Providing risk assessments in such a format to prioritise Critical Care Outreach work is a good idea |
|  | ATT2 I am positive toward the idea of assessing patient deterioration risk in such a fashion |
| BI | BI1 Were this system offered in my practice, I believe I would be a frequent user |
|  | BI2 Were this system available, but not offered in my practice, I would advocate for its use |
| PEOU | PEOU1 It would be easy for me to use this application in my clinical practice |
|  | PEOU2 It would be easy to become skillful at using this application |
|  | PEOU3 It is easy for me to understand the patient deterioration risk assessments that are presented by this application |
|  | PEOU4 I feel confident that I could find the information I wanted in this application |
| PU | PU1 Death and unplanned ICU admission risk assessments presented in this manner would enhance the effectiveness of the Critical Care Outreach team |
|  | PU2 This application would make it easier to prioritise Critical Care Outreach resources |
|  | PU3 This application would be useful to the Critical Care Outreach team |
| Free-text | Do you have any additional comments that you would like to make regarding the clinical utility of the proposed tool? |

#### 3.3.2. Effect of Role

*H6* is supported, as there is a statistically significant difference between the perceived usefulness between nurses and non-nurse respondents. A more meaningful inter-group comparison for this hypothesis would be nursing staff compared to medical staff, as there would be more uniformity in the scope of the roles being analysed, however once again we are limited by the small data set.

#### 3.3.3. Effect of Workload

There is no statistically significant difference in the PU latent factor when comparing the group of respondents who spend all or the majority of their working week assigned to relevant clinical patient deterioration tasks, versus those who spend only part of their time or ad-hoc assignment in this role, so *H7* is not supported.

### 3.4. Free-text responses

23 subjects provided input to the optional free-text comment at the end of the study. These responses were typically very short (mean 175 characters, s.d. 144), but despite this, consistent themes were strongly evident. Only three comments were too general in their nature to fit into at least one of the identified themes. We report here the results of this abstraction for the purposes of driving the direction of future exploratory analyses ^3^. The most common response type (10) referred to a specific setting (physical or logical) — either in support of the utility of this tool in a given context, or where there was some setting-specific limitation in its use.

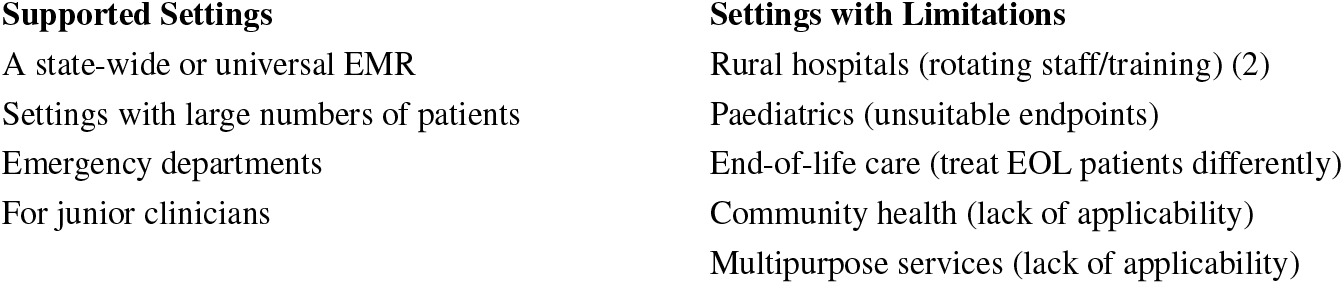

Four responses referred to the impact of this tool on quality of care, two of which repeated the same concern that such automation must not be allowed to impact or supersede face to face care. The other two were more positive with respect to the potential effect that the tool may have on patient outcomes by introducing timely and specific alerts.

Workload was also mentioned in four responses, specifically: that any manual steps will increase the load of already overburdened critical care teams (2 responses); the concern that availability must be straightforward and flexible for it to be useful; and that a formalisation such as this may generate data to support implementing dedicated response teams where they do not already exist.

Three subjects commented on the value of synthesising large volumes of data from numerous sources, and the additional benefit that this may provide in terms of a highlevel overview of current acuity levels.

Finally, three subjects provided general caveats or considerations that they considered key to the successful implementation of this tool. These were: for this to be useful, it must be possible to know the reason behind the deterioration; the necessity of specialist clinician informatician involvement, particularly with respect to privacy and security; and the importance of a carefully designed roll-out phase.

## 4. Discussion

### 4.1. Study Measure

This study demonstrated the applicability of the TAM measures to describe the attitudes and intentions of clinicians to adopt the proposed AI system for the purpose of decision support in a patient deterioration context. Although it was not possible to fully validate the ATT and BI measures, the retained relationships between latent factors were consistent with prior literature, and explained a large proportion of the variance in the overall opinions of the target users.

It is possible that the need to merge the BI and ATT factors was due to the composition of the study population, which is over-representative of very senior clinicians. BI items (when interpreted in their literal sense) ask a subject to reflect on the likelihood of an action on their part that may fall outside of the scope of a managerial role (e.g. *I would be a frequent user*), and thus breaks the directness of translation of a positive overall attitude into a behavioural intention to use. In future studies, it would be illustrative to identify the role of decision-maker as distinct from likely user, and adjust the BI items to account for this difference in remit.

### 4.2. Clinician Readiness

As seen in Table 5, there is an overall positive view of all the latent factors in the research model. This means that a subject is more likely than not to perceive that the system under review is both somewhat useful and somewhat easy to use, although this is not universally true. The readiness to adopt this technology would be best described as ‘moderate’, with nurses somewhat more likely than other clinicians to have a positive view of its potential utility.

The favourable view of nurses as to the usefulness of this system is a good indicator of support for a pilot implementation, as nurses are generally more burdened with administrative tasks introduced by hospital information management systems. It would be reasonable therefore to expect them to be a more skeptical user group for novel hospital IT programs. Their positive assessment should be taken as evidence that this system has potential to fit in well with this workflow, and that as a user group they are open to the idea of automated information synthesis and risk assessments that could augment their patient care. There is also evidence that nurses do not always find the patient care escalation process to be without friction when based on intuition alone [46], so it would be informative to explore in what capacity nursing staff perceive this system to be useful - whether for its information-synthesis capacity, risk assessment, or as an additional measure that can be used to make the case for patient prioritisation.

The effects of the PU and PEOU factors upon the combined BI/ATT endogenous factor are fairly equally weighted in this model, which goes against *H3b*. This may be due to the novelty of this system, where it is easier for a user to assess ease of use against their mental models of existing clinical software than it is for them to fully imagine how it will assist their practice. Based on experience, however, we would posit that in order to take this system from the research to the clinical realm PU will in fact be a far higher barrier to cross. This was evident in the focus of the free-text responses on specific settings where the system would be most useful or where it may have limitations to its usefulness. The hypothetical nature of this system may require the introduction of an additional factor to capture the friction between a user’s general positive attitude towards the use of a system and the behavioural intention to not only use a system but also to overcoming technical and procedural changes necessary for its implementation.

Relationships between the latent factors were consistent between study groups, both in scale and factor loadings, although the variance showed some differences. More data would be required in order to further investigate these differences in variance and to infer anything about the patterns of which individual measures showed a statistically significant inter-group variation. This may also be an outcome of the seniority bias evident in the study sample.

### 4.3. Limitations

The most obvious limitation of this study is in its small sample size. We chose to prioritise the relevance and expertise of the subjects, at the expense of the available population. The results here are therefore challenging to generalise, although they give a solid basis upon which to build.

In addition, the novelty of the target system makes it difficult for subjects to meaningfully evaluate in this limited context. This is seen in the relatively high percentage of respondents who filled in the demography measures completely, but did not proceed with the questionnaire after reviewing the prototype application. Until a controlled experiment demonstrating this system in practice (or better: a working prototype) is available, any judgements of PU in particular will be insufficient to draw broad conclusions.

### 4.4. Future Work

The free-text responses were illustrative of the general concerns and objectives of this group of potential decision-makers and users, however they were insufficiently formal to draw any significant conclusions. A semi-structured interview format would be best to further explore these themes, in order to identify the specific barriers between the moderate readiness identified in this work and an actual pilot implementation.

## 5. Conclusion

Clinicians were found to be moderately favourable towards the AI decision support system that was presented as a potential prototype for the support of managing critical care outreach workloads. Nurses were somewhat more likely than other clinicians to perceive the system as useful in their practice.

## Data Availability

Raw survey data are not available due to restrictions from the relevant HREC approval.

## Appendix

### Text presented to survey respondents

#### Background

This application has been developed in order to assist the work of the Critical Care Outreach team by presenting assessments of patient deterioration risk over time.

You will be presented with a system mock-up and text describing the intended usecases.

The purpose of the questions that follow is to determine the potential usefulness, ease of use and attitudes towards such an application. In responding to these questions, please assume that the risk assessments presented have been validated to the following level of accuracy for in-patient deaths and unplanned ICU admissions.

### Application Overview

#### Data sources

The watch-list is envisaged as an application that can be run automatically, from data that is available in the clinical record in real time, i.e. pathology orders and results, medication administration records, surgical theatre bookings, ward movements and administrative data.

This list will update regularly as new data is available, producing a risk assessment for each patient.

#### Prediction of in-patient death

The prototype system is able to predict death within 24 hours with an accuracy that compares favourably to the commonly used early warning score NEWS (AUROC 0.93 vs 0.89).

#### Prediction of unplanned ICU admission

This system is able to predict unplanned ICU admission within 24 hours (defined here as admissions to ICU from any source other than direct from surgical theatres) with an AUROC of 0.77.

This accuracy and clinical applicability will be tested and reported upon separately. Your responses should focus on the usability of the application itself, assuming the accuracy described above.

Note that the prototype system does not make use of vital signs data or clinical notes, and accuracy is expected to increase significantly (particularly in regards to unplanned ICU admission) when this data is made available.

### Application Usage

(A) Summary panel -see high-level view of cases allowing an immediate overview of the risk profile of the hospital

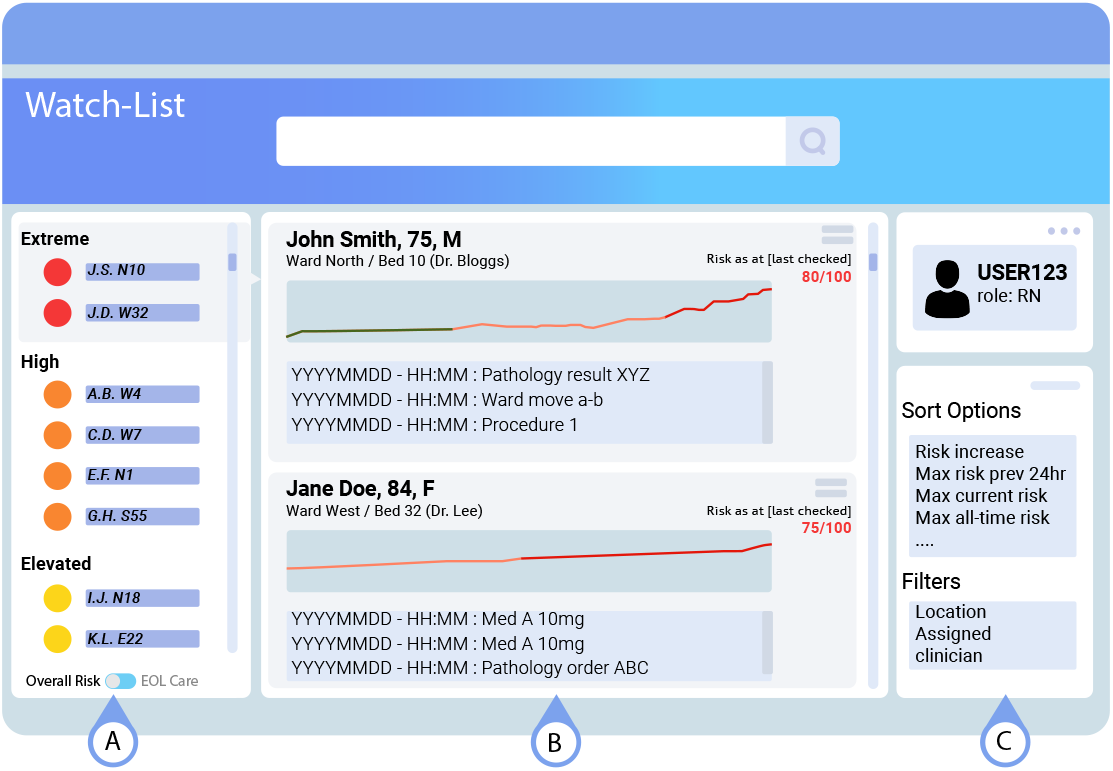

(B) Detail view -see predictions for individual cases over time to understand trends, and highlight events co-occurring with risk changes over time

(C) Control panel -sort and summarise by risk category, location, assigned clinician or other relevant parameters

**Figure 2.**
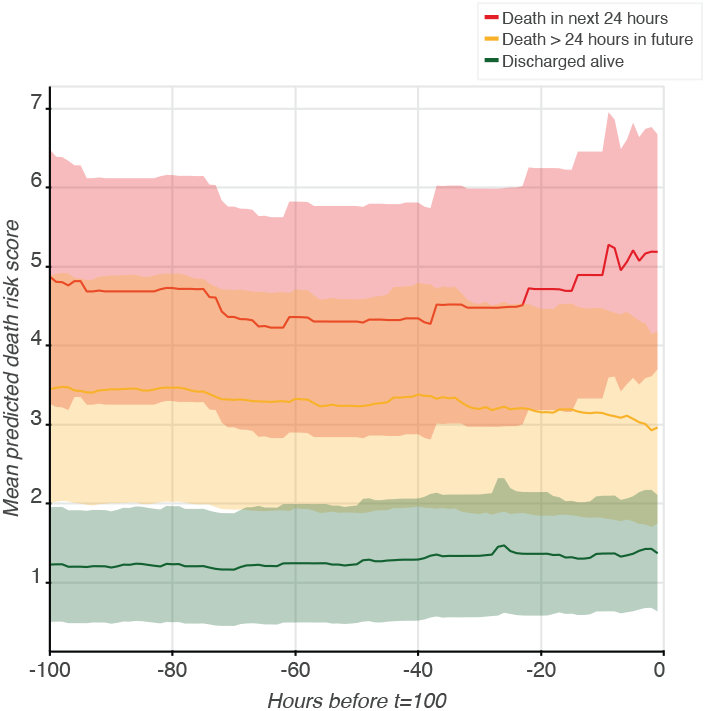
Risk changes up to prediction time.

It is not possible to give a real example for the detail panel due to privacy restrictions on the source data, so here we present the average risk trend for the first 100 hours of admission for admissions *>* 100 hours in length in the test set (grouped by actual outcome).

### Application Use Cases

A non-exhaustive list of expected use-cases includes:

1. Using the control panel to sort by location and risk, the critical care outreach nurse begins their shift and is able to rapidly assess where to visit first.
2. A medical officer is handing over at the end of their shift. They filter by assigned clinician and/or location and use the output as a priority list to work through for the handover conversation.
3. A rapid-response is called on a patient. The responding medical officer is not familiar with this patient, however they are able to use the view of risk over time, together with the listed events summarised below, to help them get up to speed on this case more rapidly.
4. Clinicians can manually override a high-risk prediction in the instance that the patient in question is receiving end-of-life care.
5. Post-hoc analysis of outlier predictions can be used to classify and identify unexpected deterioration in order to drive institution-level policies.

## Footnotes

2 Note that these examples are limited to those demonstrating successful implementation and real-world use, as opposed to studies describing model development, which are plentiful.

3 Note that it is outside of the bounds of the ethical approval of this study to report any quotes directly.

